# Genome-wide polygenic risk score method for diabetic kidney disease in patients with type 2 diabetes

**DOI:** 10.1101/2021.07.09.21260114

**Authors:** Ha My T. Vy, Sergio Dellepiane, Kumardeep Chaudhary, Alexander Blair, Benjamin S Glicksberg, Steven G Coca, Lili Chan, John Cijiang He, Ron Do, Girish N Nadkarni

## Abstract

Diabetic kidney disease (DKD) is considered partially hereditary, but the genetic factors underlying disease remain largely unknown. A key barrier to our understanding stems from its heterogeneity, and likely polygenic etiology. Proteinuric and non-proteinuric DKD are two sub-classes of DKD, defined by high urinary albumin-to-creatinine ratio (UACR) and low creatinine estimated glomerular filtration rate (eGFR). Prior genome-wide association studies (GWAS) have identified multiple loci associated with eGFR and UACR. We aimed to combine summary statistics from previous GWAS’ for eGFR and UACR in one prediction model and associate it with DKD prevalence. We then tested this using genetic data from 18,841 individuals diagnosed with type 2 diabetes in UK Biobank. We computed two genome-wide polygenic risk scores (GPS) aggregating effects of common variants associated with the two measurements, eGFR and UACR. We show that including both GPS’ in a single model confers significant improvement in comparison with the single GPS model generated from GWAS summary statistics for DKD. We also find in replication analysis in 5,389 individuals in the multi-ethnic Bio*Me* Biobank, that although the combined model had consistent direction of association, the lowest performance was in individuals with recent African ancestry. In summary, we show that joint modeling of polygenic associations of eGFR and UACR is more significantly associated with DKD than individual modeling as well as a GPS comprised of only DKD summary statistics and may be used to gain insights into biology and progression. However, efforts should be made to develop and validate polygenic approaches in diverse populations.

## INTRODUCTION

Diabetes is the most common cause of kidney disease in the United States.^1^ Approximately 25-40% of patients with type 2 diabetes (T2D) will develop kidney disease-termed as diabetic kidney disease (DKD).^2^ The incidence of DKD is increasing, and thus better approaches to understanding, risk stratifying and prognosticating DKD are needed.^3,4^

DKD is a heritable disease as indicated by different incidence in certain groups, familial aggregation and a significant heritability index. ^5, 6,7^. These point towards a relevant genetic component of DKD; however, no single mutation or gene has been identified to have a large impact on disease development. Thus, progress towards understanding DKD at a genetic level will need to consider the polygenic nature of this complex disease.

A genome-wide polygenic risk score (GPS), an index that aggregates the cumulative effects of millions of common variants across the genome, has ability to identifies individuals with high risk for diseases with polygenic pathogenesis.^8^ In several multifactorial disorders (e.g. coronary artery disease, atrial fibrillation, and type 2 diabetes) GPS scores are as predictive of disease incidence as Mendelian mutations, but can identify much larger groups of individuals.^8^ However, unlike other polygenic diseases, efforts in using GPS to estimate DKD risk have shown very limited success.^9^ The cardinal features of DKD are urinary albumin-to-creatinine ratio (UACR) ≥30mg/g or estimated glomerular filtration rate (eGFR) <60ml/min/1.73m^2^ in patients with diabetes and without other causes of renal injury.^10^ While albuminuria was once considered a key feature of DKD; up to 30% of DKD patients with disease progression do not have proteinuria and do not respond to antiproteinuric therapy,^1^ suggesting distinct disease pathways and, likely, different genetic causes. As such, several authors prefer to distinguish between proteinuric (pDKD) and non proteinuric DKD (npDKD).

However, the categorization of DKD based upon a threshold for eGFR and/or UACR, combined with the small sample size of available studies, has resulted in small effect sizes for the majority of risk variants reported for DKD.^11-14^ In contrast, GWAS for eGFR and UACR, as continuous traits, have been conducted for much larger samples and multiple significant associations with modest effect sizes have been uncovered.^15,16^

We hypothesized that incorporating genetic information from those studies and jointly incorporating GPS from both eGFR and UACR may improve the association with DKD. We developed a model combining information from two GPSs for two relevant quantitative phenotypes, eGFR and UACR in individuals with T2D and tested the performance of these scores using the UK Biobank (UKBB), a predominantly European cohort, and further evaluated the performance of the model in different ethnicities using Bio*Me* Biobank, a diverse clinical biobank cohort in the United States. Finally, we compared the association of this joint modeling approach vs. that of model consisting of only GWAS significant hits for DKD.

## METHODS

The overall study workflow is shown in **Figure 1**.

**Figure 1.**
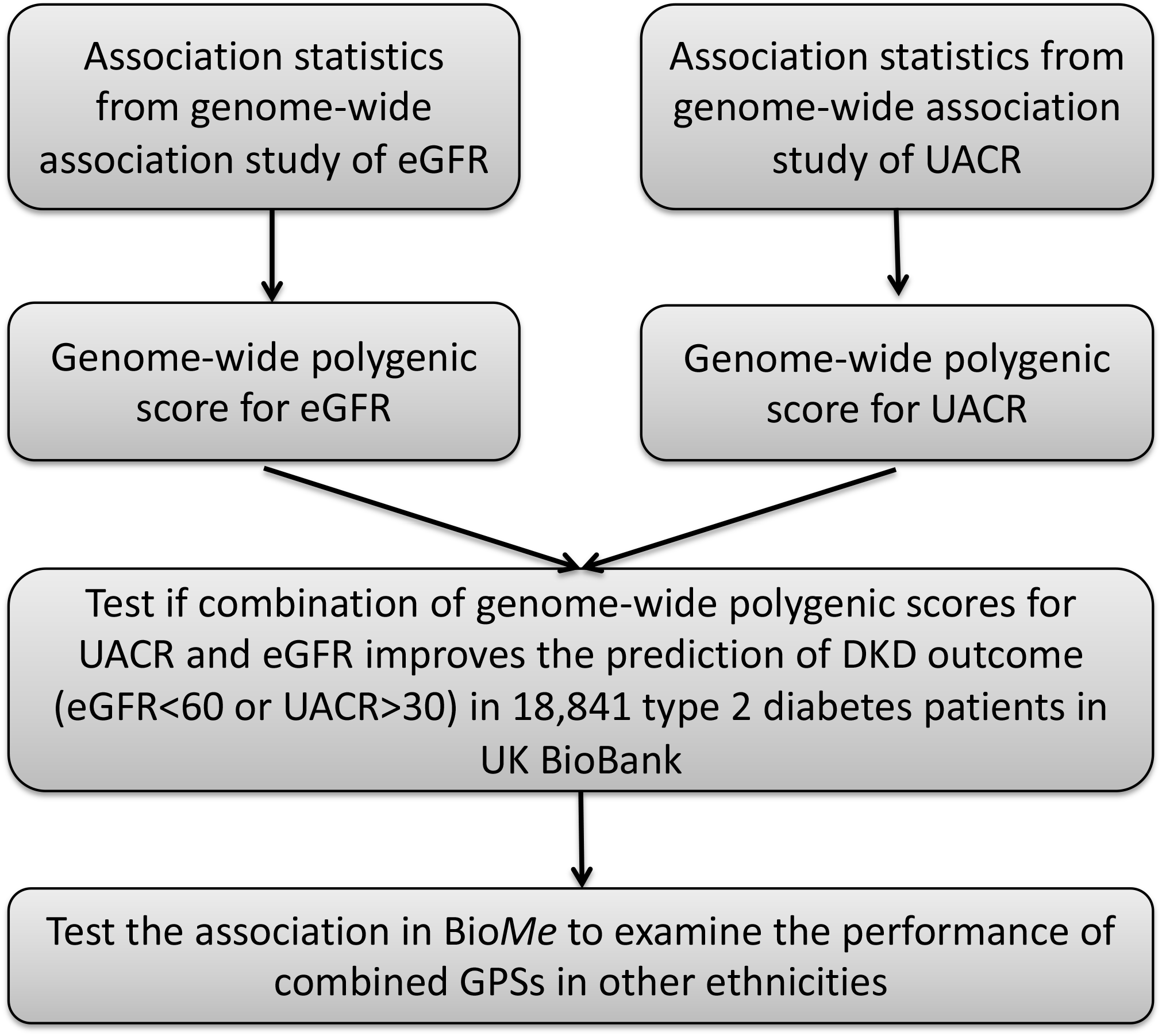
Overall Workflow and Flowchart for the Study. Using two recent GWASs for two phenotypes, eGFR and UACR, two independent GPSs were derived. A prediction model of DKD incorporating the two GPSs was then tested in 18,841 individuals of European ancestry with T2D from UKBB and subsequently validated in other ethnicities using Bio*Me* Biobank.

### Description of Cohorts

The UKBB is a large prospective population-based cohort study of approximately 500,000 individuals aged between 39-73 who were recruited from 22 assessment centers across the United Kingdom between 2006-2010.^17,18^ Genotyping was performed on the Affymetrix Axiom UK Biobank array (∼450,000 individuals) or the UK BiLEVE array (∼50,000 individuals) and then imputed using the Haplotype Reference Consortium (HRC) combined with UK10K haplotype resource. 84% of the cohort (409,703 individuals) is of European ancestry, distributed across the United Kingdom.

The BioMe Biobank is an EHR-linked clinical care biobank cohort comprised of over 45,000 participants from diverse ancestries (African, Hispanic/Latino, European and Other ancestries), with accompanying genome-wide genotyping data for 32,595 participants. Along with the genetic information, Bio*Me* is linked to a wide array of biomedical traits, originating from Mount Sinai’s system-wide electronic health record (EHR). Enrollment of participants is predominantly through ambulatory care practices; and as a whole are representative of the larger patient population served by the Mount Sinai Health System.^2^ Genotyping data of 32,595 participants was performed on the Illumina Global Screening Array (GSA) platform. Quality control and imputation of genotyping data are described in the supplementary appendix.

### Identification of T2D case/control status

Statistical analyses in this study were restricted to patients with T2D. In UKBB, T2D was defined by International Classification of Diseases (ICD-9 250.00, 250.10; ICD-10 E11) codes or self-reported illness codes (1223) through verbal interview. In Bio*Me*, T2D was identified based on the case and control selection algorithm developed by the eMERGE consortium (https://emerge-network.org/) which takes into account ICD codes for T1D and T2D, lab values, and physician entered diagnosis.

### Statistical Analyses

#### a. Generation of GPSs

GPS measures the cumulative impact of common variants on the risk of certain disorders or diseases. We assumed the impact of independent variants to be additive. Thus, for each individual, we computed GPS by taking the sum of the dosage of the effective allele at each single nucleotide polymorphic site (SNP) weighted by their effect on the outcome under consideration.

For eGFR, weights were assigned based on GWAS summary statistics generated from 113,814 European ancestry (EA) individuals.^15^ For UACR, GWAS summary statistics generated from 54,450 European ancestry individuals^16^ was used. A GPS for DKD was also generated for comparison, using summary statistic from a GWAS on 5,717 samples.^14^ To account for linkage disequilibrium, we implemented the LDpred computational algorithm^19^ to generate adjusted GWAS summary statistics based on a linkage disequilibrium reference panel of 503 Europeans from 1000 Genome phase 3 version 5. For each phenotype, nine adjusted GWAS summary statistics corresponding to nine given values of the fraction of causal variants (ρ = 1.0, 0.3, 0.1, 0.03, 0.01, 0.003, 0.001, 0.0003, and 0.0001) were generated from the original GWAS. A candidate polygenic score was then generated for each adjusted GWAS using the PRSice software^20^ with an option to remove all polymorphic sites with ambiguous strands (A/T or C/G).

#### b. Testing of GPSs

The associations between npDKD, pDKD, and corresponding GPSs were tested using a simple logistic regression model adjusted for age, sex, assessment center, genotype measurement batch, and the first ten genetic principal components (PCs). Out of the nine candidate GPSs for each phenotype, the score with the strongest odds ratio and the corresponding lowest P-value was chosen for subsequent analysis.

#### c. Association of GPS_eGFR_ and GPS_UACR_ with composite DKD

Composite DKD cases were defined as individuals with either eGFR <60 mL/min/1.73m^2^ and/or UACR ≥30 mg/g. The association between composite DKD and GPSs were tested using a single polygenic score model which tested for each GPS separately, and a multi-polygenic score model, which combined two polygenic scores into one logistic regression model. Covariates included age, sex, BMI, systolic blood pressure (SBP), assessment center, genotype measurement batch and the first 10 PCs.

#### d. Validation in multiple ethnicities

Two genome-wide polygenic score, GPS_eGFR_ and GPS_UACR_, were generated for each individual in the Bio*Me* data set using the same GWAS summary statistics used for computing GPSs in UKBB. The associations between GPSs and composite DKD were then tested in European American, Hispanic American, and African American cohorts separately using a multi-polygenic score model adjusted for age, sex, SBP, BMI and the first ten PCs.

## RESULTS

Of the 18,841 individuals with T2D in UKBB, 3,498 cases (20%) had DKD. Of those with DKD, 781 cases had renal dysfunction (eGFR <60 ml/min/1.73 m^2^) without proteinuria (UACR <30 mg/g), and 2,717 cases had proteinuria (UACR ≥30 mg/g). Individuals with DKD had higher age, BMI, and SBP than individuals without DKD on average (**Table 1**).

**Table 1.** Baseline characteristics of 18,841 participants with T2D in UK Biobank cohort

| Characteristic | All Participants<br>with T2D | Participants with<br>DKD | Participants<br>without DKD |
| --- | --- | --- | --- |
| **Sample size** | 18841 | 3498 | 13889 |
| **Age in years** | 60.45 (6.73) | 61.48 (6.50) | 60.21(6.76) |
| Female | 7111 (38) | 1250 (36) | 5245 (38) |
| **Creatinine (umol/L)** | 76 (26) | 92 (49) | 73 (14) |
| **Microalbumin (mg/L)** | 36 (169) | 143 (369) | 11 (8.5) |
| **Body Mass Index in**<br>**kg/m<sup>2</sup>** | 32 (6) | 32 (6) | 31 (5.6) |
| **Systolic Blood**<br>**Pressure in mm of Hg** | 145 (19) | 148 (22) | 144 (18.26) |

All categorical values are in n (%) and continuous values are in mean (SD)

### Performance of each GPS

GPS_eGFR_ corresponding to 1% of causal variants (ρ=0.01) was the best predictor of npDKD (p-value=3.33×10^−26^; R^2^=0.07) (**Table S1**). For pDKD, GPS_UACR_ corresponding to 0.3% of causal variants (ρ=0.003) had the best performance (p-value=1.8×10^−4^ ; R^2^ = 0.006) (**Table S2**). We therefore brought forward these two GPSs for inclusion in the multi-polygenic score model.

We observed that increased GPS was associated with increased risk of DKD (**Figure 2**). The prevalence of DKD increased proportionally with GPS_eGFR_ and GPS_UACR_, with an approximate 7% increase in npDKD from the lowest decile to the highest decile of GPS_eGFR_ (**Figure 2A**) and 5% increase in pDKD from the lowest decile to the highest decile of GPS_UACR_ (**Figure 2B**). The median of GPS decile score was 6 for individuals with DKD cases versus 5 for individuals without DKD (**Figure 2C** and **D**).

**Figure 2.**
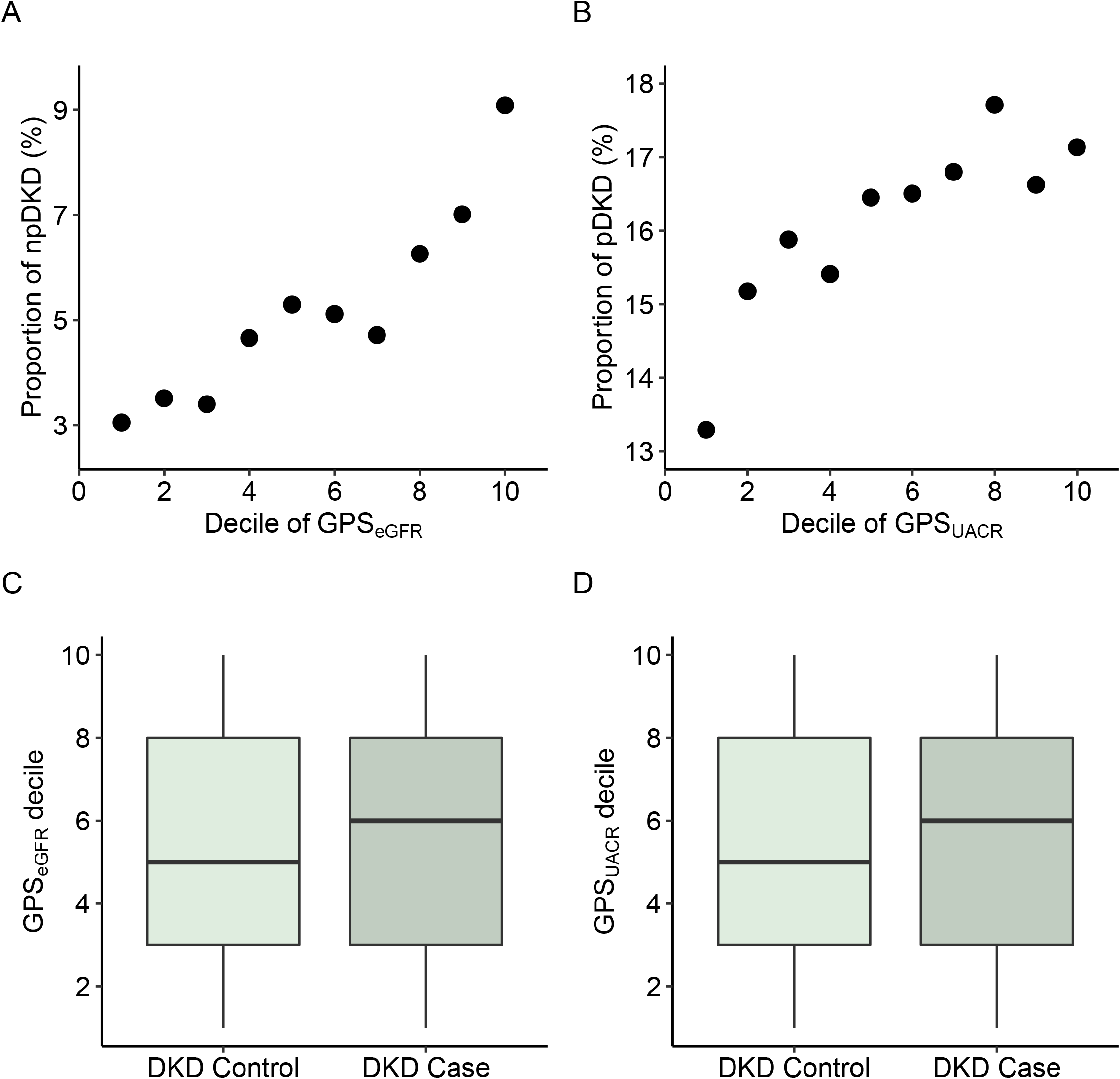
Association with prevalent DKD increases with increasing genome-wide polygenic scores. Top row: proportion of DKD incidents in each GPS decile. npDKD is defined by eGFR <6060ml/min/1.73m^2^ (A); pDKD is defined by UACR<30mg/g (B). Bottom row: GPS decile among DKD cases versus controls. Each boxplot reflects the decile range from first to third quartiles and the horizontal line in the middle indicates decile mean.

### Joint prediction of GPS_eGFR_ and GPS_UACR_

Although the two GPSs are independent (Pearson correlation (GPS_eGFR_, GPS_UACR_) = -0.05), results from logistic regression test using multi polygenic score model showed strong associations between both GPSs and DKD (p-value 1.1×10^−8^ and 1.5×10^−4^ for GPS_eGFR_ and GPS_UACR_ respectively; **Table 2**). The contribution of each GPS to DKD risk stratification was modest, evidenced by odds ratio of 1.13 for GPS_eGFR_ and 1.08 for GPS_UACR_. However, in comparison with the single GPS model which uses GPS generated from GWAS summary statistic for DKD (**Table 2**), the multi-polygenic score model showed a significant improvement.

**Table 2.** Performance of joint GPS model and single GPS model in predicting DKD.

| Model | Predictor | Odds ratio | 2.5% CI | 97.5% CI | P-value | $R^2$ |
| --- | --- | --- | --- | --- | --- | --- |
| Joint GPS | $\text{GPS}_{\text{eGFR}}$ | 1.13 | 1.09 | 1.18 | $1.08 \times 10^{-8}$ | 0.22 |
| | $\text{GPS}_{\text{UACR}}$ | 1.08 | 1.04 | 1.13 | $1.47 \times 10^{-4}$ | |
| **Single GPS** | **GPS<sub>DKD</sub>** | 1.06 | 1.02 | 1.10 | 0.004 | 0.19 |

We next explored whether there was any significant clinical difference between individuals at the high extreme distribution of polygenic score distributions compared to individuals at the low extreme distribution. We observed an increasing strength of association between higher polygenic scores and DKD compared to those with lower polygenic scores (**Table 3**). A particularly strong enrichment in DKD was observed in individuals belonging to the highest 10% of both GPS_eGFR_ and GPS_UACR_ distributions, with > two-fold adjusted odds of DKD diagnosis (OR 2.15, 95% CI 1.02-4.68; p=0.04).

**Table 3.** Joint GPSs stratification analysis

| Sample grouping based on GPSs | OR | 2.5% CI | 97.5% CI | P-value |
| --- | --- | --- | --- | --- |
| **Top 50% vs bottom 50%** | 1.35 | 1.21 | 1.51 | $1.9 \times 10^{-7}$ |
| **Top 40% vs bottom 40%** | 1.45 | 1.26 | 1.68 | $3.5 \times 10^{-7}$ |
| **Top 30% vs bottom 30%** | 1.68 | 1.38 | 2.06 | $3.1 \times 10^{-7}$ |
| **Top 20% vs bottom 20%** | 1.69 | 1.24 | 2.31 | $8.7 \times 10^{-4}$ |
| **Top 10% vs bottom 10%** | 2.15 | 1.02 | 4.68 | $4.7 \times 10^{-2}$ |

### Performance of GPS_eGFR_ and GPS_UACR_ in different ethnic groups

We assessed the performance of the multi polygenic score model in three different ethnic groups in Bio*Me*: 1,855 African American (AA); 2,684 Hispanic American (HA); and 850 European American (EA, **Table 4**). Logistic regression tests between DKD and GPSs showed only one significant association between DKD and GPS_eGFR_ in the HA group, possibly because this is the group with largest sample size, however, we observed consistent direction of association (OR>1). Observed R^2^ is highest for EA group and lowest for AA group, indicating that our GPSs perform better in European and European admixed ethnicities.

**Table 4.**
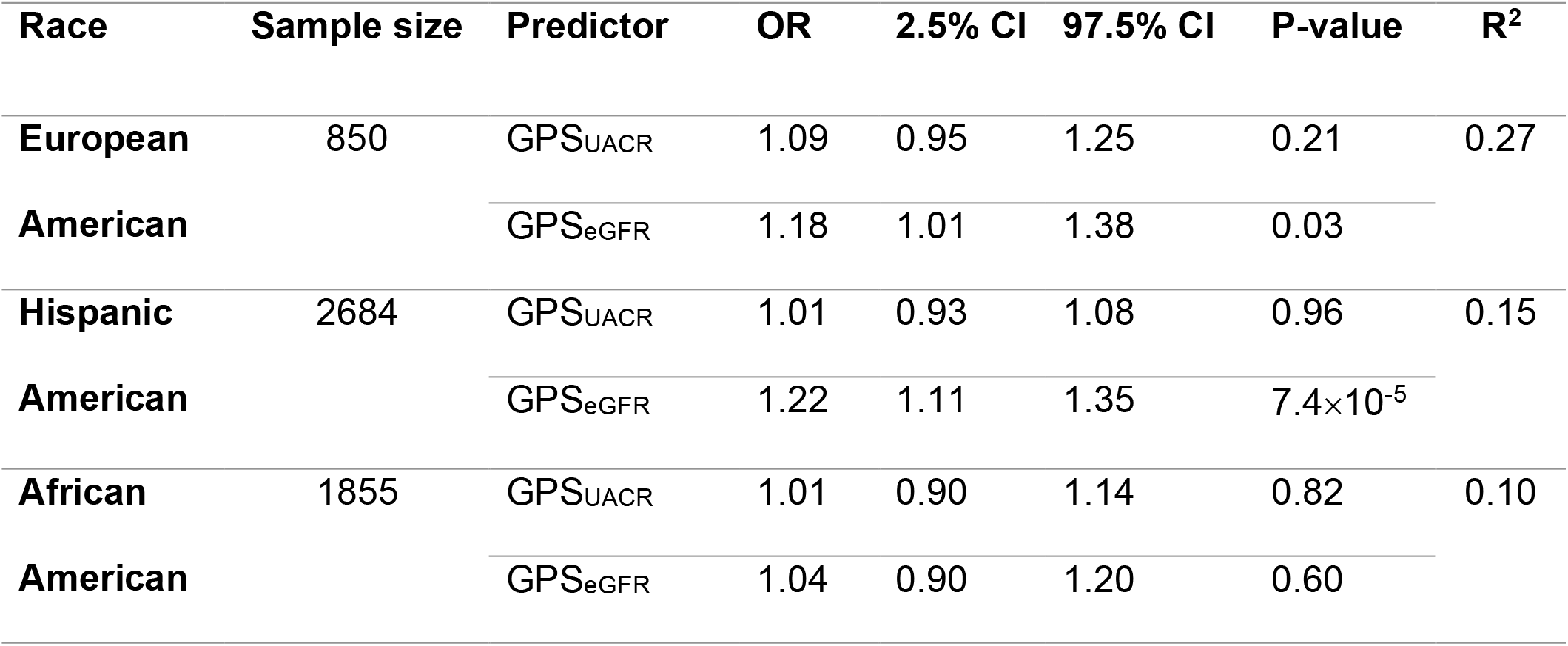
Performance of the two GPSs in multiple ethnic groups

## DISCUSSION

In this study we developed a polygenic approach which combined genetic information from two genome-wide polygenic information from two DKD defining traits-UACR and eGFR. First, we show that in UKBB patients with T2D, individuals belonging to the top deciles of the two GPSs were at more than two-fold increased odds of DKD relative to those in the bottom deciles. Second, we show that combining information from both eGFR and UACR, has superior association with DKD compared to using GWAS significant hits from DKD alone. Finally, we show that the performance of this joint modeling is significantly worse in African Americans compared to European Americans, highlighting the need for improved diversity in these studies.

Although GPS have been developed and validated for many complex diseases including T2D^21,22^ and chronic kidney disease (CKD),^23,24^ previously, only one study has been published that generated and validated GPS for kidney disease in T2D. Liao et al 2019^9^ used a polygenic score in Han Chinese patients with T2D and found that adding genetic factors to the clinical factors can improve risk prediction, however their GPS was comprised of only seven DKD susceptibility SNPs with a sample size of less than 1000 individuals. This study, on the other hand, explored the summed effect of millions of SNPs. Moreover, instead of using one single GPS, we utilized genetic information from two independent GPSs for cardinal quantitative traits of DKD (eGFR and UACR)

Both reduced eGFR and albuminuria are clinical markers of diabetic nephropathy and have a distinctive underlying genetic basis. Clinically significant albuminuria is prevalent in certain populations with diabetes, and progression to higher levels of albumin loss in the urine varies among ethnicities.^25,26^ In addition, albuminuria is highly heritable among families with diabetes and hypertension,^27^ and progressive albuminuria is also associated with significant reductions in eGFR. Similarly, eGFR decline is heritable^28^ and variations in glomerular filtration is a risk factor for CKD among diabetics.^29^ Previous studies have shown that combination of changes in both eGFR and UACR is a better predictor of major kidney events than when the two are assessed separately.^30^ Our study further demonstrates that combining genetic information from two independent GPSs for the two quantitative traits associates with DKD more strongly than just using genome wide significant hits for DKD (**Table 2**).

The risk for developing DKD has been shown to be partially due to genetic factors, yet we know very little about its genetic architecture. Although a major effort has been made to discover variants and genes with large effect on DKD in type 1 or type 2 diabetes, only a few variants, without robust statistical power or independent validation, have been reported.^31^ While the lack of compelling evidence for large effect mutation and gene is attributable to small number of patients in those studies, it also suggests that DKD is likely polygenic and patient susceptibility may result from the summative effects of numerous common variants with individually small effects. Associations of GPSs with DKD in our study support this argument, as the optimal GPS corresponds to a fraction of 1% (∼18,000) causal variants for npDKD and 0.3% (∼5,400) causal variants for pDKD (**Table S1-2**). In fact, GWAS studies for eGFR and UACR have discovered some of the genome-wide significance variants to be part of biologically interesting pathways which include and are not limited to glucose homeostasis, regulatory function in the kidney, reduced reuptake of albumin, decreased vesicular trafficking of albumin and alteration of glomerular basement membrane proteins.^32-35^ Moreover, kidney dysfunction in diabetes may be a result of not only primary injury to the nephron from hyperglycemia, but also as a consequence of secondary processes such as age, and systolic blood pressure. These biological pathways are complex and comprise thousands of SNPs, suggesting a future direction of DKD studies to focus on the polygenic nature of the disease and investigate pathways regulated by multiple genetic variants.

There are several limitations in this study. First, it is unclear whether the subset of patients identified with higher polygenic risk scores will develop more severe DKD, as longitudinal data is currently not publicly available from the databases used. Second, how GPS performs compared to traditional risk factors in DKD prediction is still to be determined and may require a more longitudinal study before disease onset. Third, replication analysis in varying ethnicities in the Bio*Me* database, particularly AA and HA yielded poor results, suggesting that further studies with larger sample size and GWAS conducted for samples from different ethnicities would be necessary to generate more statistically robust validation.

Regardless of these limitations, this study serves as a proof of principle that jointly modeling polygenic contributions of eGFR and UACR can improve association with DKD. It underscores the polygenic nature of DKD and verifies that genetic information accumulated in GPSs is an important risk predictor that could be implemented in prediction models.

## Data Availability

The authors confirm that the data supporting the findings of this study are available within the article and its supplementary materials.

## ACKNOWLEDGMENTS

The Bio*Me* healthcare delivery cohort at Mount Sinai was founded and maintained with a generous gift from the Andrea and Charles Bronfman Philanthropies. The authors thank the individuals who were involved in the quality control and/or file handling for the exome sequencing and genome-wide genotyping data, including Aayushee Jain, Kumardeep Chaudhary, Lisheng Zhou, Michael Preuss, Quingbin Song, Stephane Wenric, and Steve Ellis. Research reported in this paper was supported by the Office of Research Infrastructure of the National Institutes of Health under award numbers S10OD018522 and S10OD026880. This study has been conducted using the UK Biobank Resource under Application Number ‘16218’. Iain S. Forrest was supported by T32GM007280, the Medical Scientist Training Program Training Grant from the National Institute of General Medical Sciences of the National Institutes of Health. Ron Do is supported by R35GM124836 from the National Institute of General Medical Sciences of the National Institutes of Health, and R01HL155915 and R01HL139865 from the National Heart, Lung, and Blood Institute of the National Institutes of Health. Girish Nadkarni is supported by R01DK127139 from the National Institute of Diabetes and Digestive and Kidney Disease and by R01HL155915 from the National Heart, Lung, and Blood Institute of the National Institutes of Health. The content is solely the responsibility of the authors and does not necessarily represent the official views of the National Institutes of Health.

## REFERENCES

1. US Renal Data System 2019 Annual Data Report: Epidemiology of Kidney Disease in the United States. Am J Kidney Dis. 2019.

2. Lindner TH, Mönks D, Wanner C, Berger M. Genetic aspects of diabetic nephropathy. Kidney International. 2003;63:S186–S191.

3. Remuzzi G, Schieppati A, Ruggenenti P. Nephropathy in patients with type 2 diabetes. New England Journal of Medicine. 2002;346(15):1145–1151.

4. Wild S, Roglic G, Green A, Sicree R, King H. Global prevalence of diabetes: estimates for the year 2000 and projections for 2030. Diabetes care. 2004;27(5):1047–1053.

5. Ma RC, Cooper ME. Genetics of Diabetic Kidney Disease-From the Worst of Nightmares to the Light of Dawn? J Am Soc Nephrol. 2017;28(2):389–393.

6. Seaquist ER, Goetz FC, Rich S, Barbosa J. Familial clustering of diabetic kidney disease. Evidence for genetic susceptibility to diabetic nephropathy. N Engl J Med. 1989;320(18):1161–1165.

7. Quinn M, Angelico MC, Warram JH, Krolewski AS. Familial factors determine the development of diabetic nephropathy in patients with IDDM. Diabetologia. 1996;39(8):940–945.

8. Khera AV, Chaffin M, Aragam KG, et al. Genome-wide polygenic scores for common diseases identify individuals with risk equivalent to monogenic mutations. Nature genetics. 2018;50(9):1219–1224.

9. Liao L-N, Li T-C, Li C-I, et al. Genetic risk score for risk prediction of diabetic nephropathy in Han Chinese type 2 diabetes patients. Scientific Reports. 2019;9(1):1–9.

10. KDOQI Clinical Practice Guideline for Diabetes and CKD: 2012 Update. Am J Kidney Dis. 2012;60(5):850–886.

11. McKnight AJ, Currie D, Patterson CC, Maxwell AP, Fogarty DG, Group WUGS. Targeted genome-wide investigation identifies novel SNPs associated with diabetic nephropathy. The HUGO journal. 2009;3(1-4):77–82.

12. Mooyaart A, Valk E, Van Es L, et al. Genetic associations in diabetic nephropathy: a meta-analysis. Diabetologia. 2011;54(3):544–553.

13. Pezzolesi MG, Poznik GD, Mychaleckyj JC, et al. Genome-wide association scan for diabetic nephropathy susceptibility genes in type 1 diabetes. Diabetes. 2009;58(6):1403– 1410.

14. Van Zuydam NR, Ahlqvist E, Sandholm N, et al. A genome-wide association study of diabetic kidney disease in subjects with type 2 diabetes. Diabetes. 2018;67(7):1414– 1427.

15. Pattaro C, Teumer A, Gorski M, et al. Genetic associations at 53 loci highlight cell types and biological pathways relevant for kidney function. Nature communications. 2016;7(1):1–19.

16. Teumer A, Tin A, Sorice R, et al. Genome-wide association studies identify genetic loci associated with albuminuria in diabetes. Diabetes. 2016;65(3):803–817.

17. Sudlow C, Gallacher J, Allen N, et al. UK biobank: an open access resource for identifying the causes of a wide range of complex diseases of middle and old age. Plos med. 2015;12(3):e1001779.

18. Bycroft C, Freeman C, Petkova D, et al. The UK Biobank resource with deep phenotyping and genomic data. Nature. 2018;562(7726):203–209.

19. Vilhjálmsson BJ, Yang J, Finucane HK, et al. Modeling linkage disequilibrium increases accuracy of polygenic risk scores. The american journal of human genetics. 2015;97(4):576–592.

20. Choi SW, O’Reilly PF. PRSice-2: Polygenic Risk Score software for biobank-scale data. GigaScience. 2019;8(7).

21. Läll K, Mägi R, Morris A, Metspalu A, Fischer K. Personalized risk prediction for type 2 diabetes: the potential of genetic risk scores. Genetics in Medicine. 2017;19(3):322–329.

22. Udler MS, McCarthy MI, Florez JC, Mahajan A. Genetic Risk Scores for Diabetes Diagnosis and Precision Medicine. Endocrine Reviews. 2019;40(6):1500–1520.

23. Fujii R, Hishida A, Nakatochi M, et al. Association of genetic risk score and chronic kidney disease in a Japanese population. Nephrology. 2019;24(6):670–673.

24. Ma J, Yang Q, Hwang S-J, Fox CS, Chu AY. Genetic risk score and risk of stage 3 chronic kidney disease. BMC nephrology. 2017;18(1):32.

25. Bryson CL, Ross HJ, Boyko EJ, Young BA. Racial and ethnic variations in albuminuria in the US Third National Health and Nutrition Examination Survey (NHANES III) population: associations with diabetes and level of CKD. Am J Kidney Dis. 2006;48(5):720–726.

26. Young BA, Katon WJ, Von Korff M, et al. Racial and ethnic differences in microalbuminuria prevalence in a diabetes population: the pathways study. J Am Soc Nephrol. 2005;16(1):219–228.

27. Mottl AK, Vupputuri S, Cole SA, et al. Linkage analysis of albuminuria. J Am Soc Nephrol. 2009;20(7):1597–1606.

28. Satko SG, Sedor JR, Iyengar SK, Freedman BI. Familial clustering of chronic kidney disease. Semin Dial. 2007;20(3):229–236.

29. Tseng CL, Lafrance JP, Lu SE, et al. Variability in estimated glomerular filtration rate values is a risk factor in chronic kidney disease progression among patients with diabetes. BMC Nephrol. 2015;16:34.

30. Ohkuma T, Jun M, Chalmers J, et al. Combination of changes in estimated GFR and albuminuria and the risk of major clinical outcomes. Clinical Journal of the American Society of Nephrology. 2019;14(6):862–872.

31. Rich SS. Genetic Contribution to Risk for Diabetic Kidney Disease. In: Am Soc Nephrol; 2018.

32. Pattaro C, Teumer A, Gorski M, et al. Genetic associations at 53 loci highlight cell types and biological pathways relevant for kidney function. Nat Commun. 2016;7:10023.

33. Teumer A, Tin A, Sorice R, et al. Genome-wide Association Studies Identify Genetic Loci Associated With Albuminuria in Diabetes. Diabetes. 2016;65(3):803–817.

34. Leng S, Bernauer AM, Hong C, et al. The A/G allele of rs16906252 predicts for MGMT methylation and is selectively silenced in premalignant lesions from smokers and in lung adenocarcinomas. Clin Cancer Res. 2011;17(7):2014–2023.

35. Devuyst O, Pattaro C. The UMOD Locus: Insights into the Pathogenesis and Prognosis of Kidney Disease. J Am Soc Nephrol. 2018;29(3):713–726.

