## Supplementary appendix for "Genome-wide polygenic risk score method for diabetic kidney disease in patients with type 2 diabetes"

**SUPPLEMENTARY METHODS**

Imputation and Quality Control:

In the UKBB, imputation was performed using the Haplotype Reference Consortium (HRC) combined with UK10K haplotype resource, leading to ~96 million variants available in the latest released imputed data version 3. We excluded 152,197 samples belonging to one of these categories: outliers in heterozygosity and missing rates, putative sex chromosome aneuploidy, self-reported non-white British ancestry, and related individuals. We excluded related individuals as one individual in each pair with relatedness up to the third degree. This led to a subsample of 335,212 individuals with available genotypic and phenotypic data.

In Bio*Me*, we performed quality control of the Global Screening Array data for N=32,595 participants and n=635,623 variants stratified by ethnicity category (African American=7794, European American=10133, Hispanic American=1172, and Other=3496). Individuals with a ethnicity-specific heterozygosity rate that surpassed +/- six standard deviations of the population-specific mean, along with individuals with a call rate of ≤95% were removed (N=684 participants in total). We then removed 84 individuals exhibiting persistent discordance between EMR recorded and genetic sex. We also removed an additional 4 individuals with phenotypically indeterminate sex and 158 duplicate individuals were also excluded from downstream analysis. In total 31,911 passed sample level QC for downstream analysis. Sites with a call rate below 95% were excluded (n=19253), along with sites that were seen to significantly violate Hardy-Weinberg equilibrium (HWE) when calculated stratified by ancestry. HWE thresholds varied by ethnicity, specifically we set a threshold of $p < 1x10^{-5}$ in African American and European American, or $p < 1x10^{-13}$ in Hispanic American (n=11,503 SNPs in total). This resulted in the retention of n=604,869 sites. Imputation of variants was performed using the 1000 Genomes Phase 3 data release as a reference panel. Genotype data which passed the above quality control filters was phased with SHAPEIT2,1 and imputed to 1000 Genomes Phase 3 reference data using IMPUTE version 2.3.2.2. Genome segments known to harbor gross chromosomal anomalies were filtered out of the final genotype probabilities files.

### SUPPLEMENTARY TABLES

**Supplementary Table 1:** Association of npDKD with $GPS_{eGFR}$ candidates generated assuming different values of the fraction of causal variants.

| Fraction of causal variants | Odds ratio | 2.5% CI | 97.5% CI | P-value | $R^2$ |
|---|---|---|---|---|---|
| 1 | 1.33 | 1.23 | 1.43 | $5.35\times10^{-14}$ | 0.059 |
| 0.3 | 1.34 | 1.24 | 1.44 | $1.08\times10^{-14}$ | 0.060 |
| 0.1 | 1.36 | 1.27 | 1.47 | $1.61\times10^{-16}$ | 0.062 |
| 0.03 | 1.43 | 1.33 | 1.54 | $3.07\times10^{-21}$ | 0.066 |
| **0.01** | **1.49** | **1.38** | **1.60** | $\mathbf{3.33\times10^{-26}}$ | **0.070** |
| 0.003 | 1.11 | 1.03 | 1.19 | $4.01\times10^{-3}$ | 0.050 |
| 0.001 | 1.02 | 0.95 | 1.10 | $6.11\times10^{-1}$ | 0.049 |
| 0.0003 | 1.11 | 1.04 | 1.19 | $3.26\times10^{-3}$ | 0.051 |
| 0.0001 | 1.13 | 1.05 | 1.21 | $1.13\times10^{-3}$ | 0.051 |

**Supplementary Table 2:** Association of  pDKD with $GPS_{UACR}$ candidates generated assuming different values of the fraction of causal variants.

| Fraction of causal variants | Odds ratio | 2.5% CI | 97.5% CI | P-value | $R^2$ |
|---|---|---|---|---|---|
| 1 | 1.08 | 1.03 | 1.12 | $3.35\times10^{-4}$ | 0.0058 |
| 0.3 | 1.08 | 1.03 | 1.12 | $3.36\times10^{-4}$ | 0.0058 |
| 0.1 | 1.08 | 1.03 | 1.12 | $3.32\times10^{-4}$ | 0.0058 |
| 0.03 | 1.08 | 1.03 | 1.12 | $3.20\times10^{-4}$ | 0.0059 |
| 0.01 | 1.08 | 1.04 | 1.12 | $2.98\times10^{-4}$ | 0.0059 |
| **0.003** | **1.08** | **1.04** | **1.13** | $\mathbf{1.81\times10^{-4}}$ | **0.0060** |
| 0.001 | 1.08 | 1.03 | 1.12 | $4.52\times10^{-4}$ | 0.0058 |
| 0.0003 | 1.06 | 1.02 | 1.10 | $6.88\times10^{-3}$ | 0.0053 |
| 0.0001 | 1.06 | 1.01 | 1.10 | $7.52\times10^{-3}$ | 0.0053 |

**Supplementary Table 3:** Association of DKD with $GPS_{DKD}$ candidates generated assuming different values of the fraction of causal variants.

| Fraction of causal variants | Odds ratio | 2.5% CI | 97.5% CI | P-value | $R^2$ |
|---|---|---|---|---|---|
| 1 | 1.06 | 1.03 | 1.10 | 0.0010 | 0.012 |
| 0.3 | 1.06 | 1.02 | 1.10 | 0.0013 | 0.012 |
| 0.1 | 1.06 | 1.02 | 1.10 | 0.0013 | 0.012 |
| 0.03 | 1.06 | 1.03 | 1.10 | 0.0011 | 0.012 |
| **0.01** | **1.07** | **1.03** | **1.11** | **0.00076** | **0.012** |
| 0.003 | 1.04 | 1.00 | 1.08 | 0.052 | 0.011 |
| 0.001 | 1.01 | 1.00 | 1.05 | 0.44 | 0.011 |
| 0.0003 | 1.01 | 0.97 | 1.05 | 0.60 | 0.011 |
| 0.0001 | 1.02 | 0.99 | 1.06 | 0.19 | 0.011 |
